# Antimicrobial resistance landscape in a metropolitan city context using open drain wastewater-based metagenomic analysis

**DOI:** 10.1101/2023.12.01.23299290

**Authors:** Manas Kumar Madhukar, Nirupama Singh, V Rajesh Iyer, Divya Tej Sowpati, Karthik Bharadwaj Tallapaka, Rakesh Kumar Mishra, Shivranjani Chandrashekhar Moharir

## Abstract

The One Health concept recognizes the inextricable interactions of the diverse ecosystems and their subsequent effect on human, animal and plant health. Antimicrobial resistance (AMR) is a major One Health concern and is predicted to cause catastrophes if appropriate measures are not implemented. In this study, to understand the AMR landscape in metropolitan city context, we performed metagenomic analysis of open drain wastewater samples. We analysed 17 samples from open drains that receive influx from human, animal, agricultural and industrial wastes. Our data suggests that macrolide antibiotics have developed the highest resistance in the city through mutations in the 23S rRNA gene, which is present in multiple pathogens including *Escherichia coli, Campylobacter jejuni, Acinetobacter baumannii, Streptococcus pneumoniae, Pseudomonas aeruginosa, Neisseria gonorrhoeae, Klebsiella pneumoniae* and *Helicobacter pylori*. Except for a few geographical locations, most other locations show a similar landscape for AMR. Considering human mobility and other similar anthropogenic activities, we suggest that such an AMR landscape may be common across other regions.

## Introduction

‘One Health’ is an integral concept recognizing the complex interactions of humans with their environment, which includes the other living components-animals, plants and microbes, and the non-living components – air and water ^1^. The non-living environmental components are continuously shared and exchanged between the living components, including the humans. Thus, it is imperative that human health is highly influenced by the shared environment and the other living organisms around them.

Environmental surveillance is an unbiased approach through which the physical components of the environment, the water and air, can be monitored for the biomarkers of human and zoonotic diseases ^2, 3^. Though the concept of one health and environmental surveillance are not new, they have recently gained specific importance during the COVID-19 pandemic, which was the most catastrophic infectious disease outbreak of global concern after the Spanish flu, in 1918 ^4, 5, 6^. Human and zoonotic pathogens, especially enteric microorganisms, find their way into the wastewater through faeces ^7^. The shotgun metagenomic analysis of the wastewater can give a comprehensive qualitative and quantitative data about the pathogens mongering amongst the communities ^8, 9, 10, 11^. Sewage treatment plants (STPs), that are common in well planned larger urban areas, usually get the wastewater influx from human communities, while open drainage systems, that are seen in rural regions, especially in developing countries, get the wastewater influx from human communities, animals, poultries, agricultural practices as well as industries. Thus, while wastewater samples from STPs can give the information specifically pertaining to the abundance, diversity and evolution of the microorganisms infecting humans, samples from open drains can serve as unbiased samples to throw light on the pathogens infecting a broader range of hosts, including animals and birds in poultries ^12^. Such an information is important in the perspective of one health, wherein the interconnectedness of various ecosystems in the environment attains recognition. Not only that, but wastewater surveillance also has the potential to be used as a complementary cost-effective approach for current infectious disease surveillance programs and can be used to observe the evolution, transmission and emergence of viruses and bacteria. During COVID pandemic, wastewater surveillance also served as an early warning system for detecting the surges in patient caseloads and identifying the new emerging variants ^13, 14^. Wastewater surveillance has thus proved its potential to be used as a warning system for future contagious disease outbreaks and can help prepare for future pandemics.

Antimicrobial resistance (AMR) is a global concern affecting human and animal health. Inappropriate use of antibiotics in the fields like agriculture, medicine, aquaculture and food industry has resulted in the wide dissemination of antimicrobial resistance across diverse habitats ^15, 16, 17, 18^. A study based on predictive statistical models estimated around 4·95 million deaths associated with bacterial AMR, including around 1·27 million deaths specifically attributable to bacterial AMR, globally in 2019 ^19^. According to the study, the six leading pathogens for deaths associated with resistance, *Escherichia coli, Staphylococcus aureus, Klebsiella pneumoniae, Streptococcus pneumoniae, Acinetobacter baumannii,* and *Pseudomonas aeruginosa*, were responsible for around 0.93 million deaths caused due to AMR and 3·57 million deaths associated with AMR in the year 2019. Another report predicted that if appropriate policy measures are not taken, an annual death toll of 10 million will be attributed to AMR by 2050 ^20^. As per the World Health Organization (WHO) report on Antimicrobial Resistance, AMR is declared as one of the top 10 global public health threats to humanity ^21^. Around 2.14 million neonatal sepsis deaths occurring globally are attributable to resistant pathogens every year ^22^. Not only that, AMR could lead to 7.5% decline in livestock by 2050 ^21, 23^. Thus, immediate attention and actions are necessary towards the geographical surveillance of AMR. Towards this, it is imperative to promote research for identifying the antimicrobial resistance genes in the pathogens in diverse niches, identifying the pathogens, the drug classes that have developed resistance and, understanding the details of the resistance mechanisms and the means of transmission between species and the disease spillover ^1^. This would aid in taking appropriate preventive measures against AMR and would help in developing new antibiotics and modify the existing ones strategically.

In this study we investigated the antimicrobial resistance landscape of an Indian metropolitan city by wastewater surveillance, using a shotgun metagenomics approach. We performed a comprehensive surveillance of 17 open drainage wastewater sites, that receive untreated sewage water from households, industries, and farming practices in the city. Our data provides a panoramic understanding about the abundance and diversity of antimicrobial resistance genes (ARGs), pathogens and resistant drug classes across the city. We also explored the predominant resistance mechanisms against the antibiotics that the pathogens have adopted in the wastewater samples that we have analysed.

## Results

### Summary of the dataset

The ‘One Health’ approach recognizes that humans do not live in isolation and that the entire ecosystem contributes towards the wholistic health of humans and animals ^24^. To understand the landscape of antimicrobial resistance at a community level, we performed shotgun metagenomic analysis of the wastewater samples collected from 17 open drainage sites from an Indian metropolitan city. The details of the sampling sites are given in Supplementary Table 1 and 12. The sequences obtained from the samples were analysed against the Comprehensive Antibiotic Resistance Database (CARD) for identifying the antimicrobial resistant ontologies (ARO), antimicrobial resistance genes (ARGs), the pathogens carrying the ARGs, the resistant drug classes and the mechanism of drug resistance ^25, 26^.

In our sample set, the average number of raw reads per sample was 33.98 million (median: 34.2; range: 23.56 – 39.85; standard deviation: 3.57) and a total of 577.57 million reads spanning 8.7 x 10^10^ nucleotides were sequenced across all the samples. Of the 577.57 million reads, 2.48 million reads (0.43%) were mapped to the CARD. Across all the samples, the average number of reads mapped per sample was 0.14 million (median: 0.16; range: 0.034-0.23; standard deviation: 0.065). The least number of mapped reads belonged to the sample GN (0.034 million) which was closely followed by the sample AA (0.043 million) and RRC (0.044 million). Of note, the number of raw reads for all the three samples were above the average, at 35 million and thus, cannot account for the lower number of mapped reads.

### Antimicrobial resistance ontology landscape

CARD uses AROs for the classification of the antimicrobial resistance genes data and the AROs are used as vocabularies for the ARGs, the associated products, and their mechanism of action^26^. Upon parsing the mapped reads, AROs were identified in the dataset. The AROs with mapped reads exhibiting more than 95% coverage were considered further for the analysis in this study ^10^. The AROs found following this criterion, accounted for a majority fraction of total mapped reads however, this fraction varied between the samples. A total of 287 unique ARO hits were observed across the samples from the 17 different locations. The 287 unique AROs led to the identification of corresponding 89 unique ARG hits, 72 unique drug class hits, 89 unique pathogens carrying ARGs hits, and 13 unique resistance mechanism hits across all samples (Figure 1A, B and Supplementary Table 1). On an average, 114.53 ARO hits per sample (median-115; range: 24-208 hits; standard deviation: 46.3) were observed in the 17 samples. The list of the AROs seen across the 17 sampling locations is given in Supplementary Table 2. The sampling location CP showed maximum number of ARO hits of 208, while the location AA showed the least number of AROs hits of 24 (Figure 1A, Supplementary Table 1 and 2).

**Figure 1:**
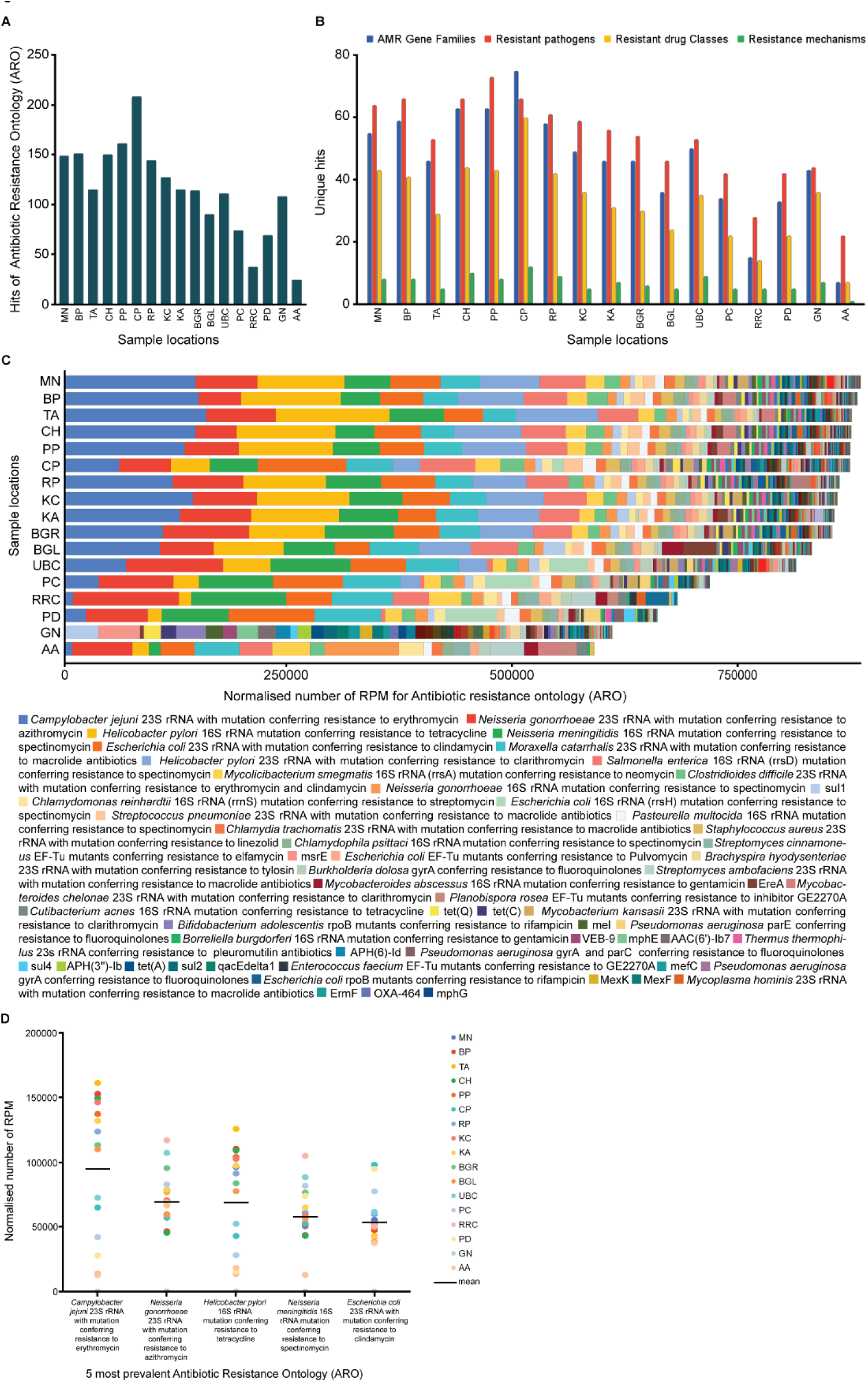
Antimicrobial resistance landscape across 17 locations in the metropolitan city. **A)** Bar diagram showing the number of hits for antimicrobial resistance ontologies (AROs) in 17 locations in the city. **B)** Bar diagram showing the number of hits for antimicrobial resistance genes (ARGs), pathogens, resistant drug classes and resistance mechanisms in the locations. **C)** Bar diagram showing the distribution pattern of the prevalence of the AROs in the 17 locations. Y axis represents the normalised reads per million of the AROs. **D)** Scatter plot showing the normalized reads per million of the 5 most prevalent AROs across the 17 locations.

While the number of ‘hits’ of AROs give information about the diversity of the AROs in a particular sample, the number of ‘reads’ of an ARO give an estimate about the prevalence of that ARO. Amongst the 17 locations, the location MN showed the highest count of normalized reads per million (RPM), followed by BP, TA, CH and PP (Figure 1C). The location AA showed the least number of normalized reads, followed by the location GN. The ARO, ‘*Campylobacter jejuni* 23S rRNA with mutation conferring resistance to erythromycin’ was the most common across the city with a prevalence of 11.8% and having an average 94879.3 RPM (Figure 1C, D, Supplementary Figure 1 and Supplementary Table 3). It was followed by ‘*Neisseria gonorrhoeae* 23S rRNA with mutation conferring resistance to azithromycin’ with a prevalence of 8.7% and mean 69509.4 RPM, ‘*Helicobacter pylori* 16S rRNA mutation conferring resistance to tetracycline’ with a prevalence of 8.6% having mean 68907 RPM, ‘*Neisseria meningitidis* 16S rRNA mutation conferring resistance to spectinomycin’ with a prevalence of 7.2% having mean 57879.5 RPM and ‘*Escherichia coli* 23S rRNA with mutation conferring resistance to clindamycin’ with a prevalence of 6.7% having mean 53592.2 RPM. These five AROs were present in all sampling sites, except for GN which showed a different ARO distribution pattern than all other sites. The most prevalent ARO in GN was msrE (Figure 1 C, D and Supplementary Table 3). MsrE, an ABC-F subfamily protein that confers resistance to erythromycin and streptogramin B antibiotics, was seen in all sampling locations except for the two locations-RRC and AA.

### ‘23S rRNA gene with mutation conferring resistance to macrolide antibiotics’ is the most prevalent ARG in the wastewater samples

89 unique ARG hits were seen in the 17 samples across the city (Supplementary Table 1 and 4). Concurrent with the counts of ARO hits, the location CP that showed the highest count of ARO hits of 208, showed the highest count of unique ARG hits of 75, amongst all the locations, suggesting the highest diversity of the ARGs in that location (Figure 2A, Supplementary Figure 2, Supplementary Table 1 and 4). The location AA showed the lowest number of ARG hits of 24 of which 7 were unique-‘23S rRNA with mutation conferring resistance to macrolide antibiotics’, ‘16s rRNA with mutation conferring resistance to aminoglycoside antibiotics’, ‘elfamycin resistant EF-Tu’, ‘16S rRNA with mutation conferring resistance to tetracycline derivatives’, ‘fluoroquinolone resistant gyrA’, ‘23S rRNA with mutation conferring resistance to lincosamide antibiotics’, ‘antibiotic-resistant rpsL’, suggesting that the location AA had the lowest diversity of the ARGs. Amongst these 7 ARGs, except for the ARG, ‘antibiotic-resistant rpsL’, which was not seen in the locations KA, GN and BGL, the other 6 ARGs were present in all other locations but not in GN.

**Figure 2:**
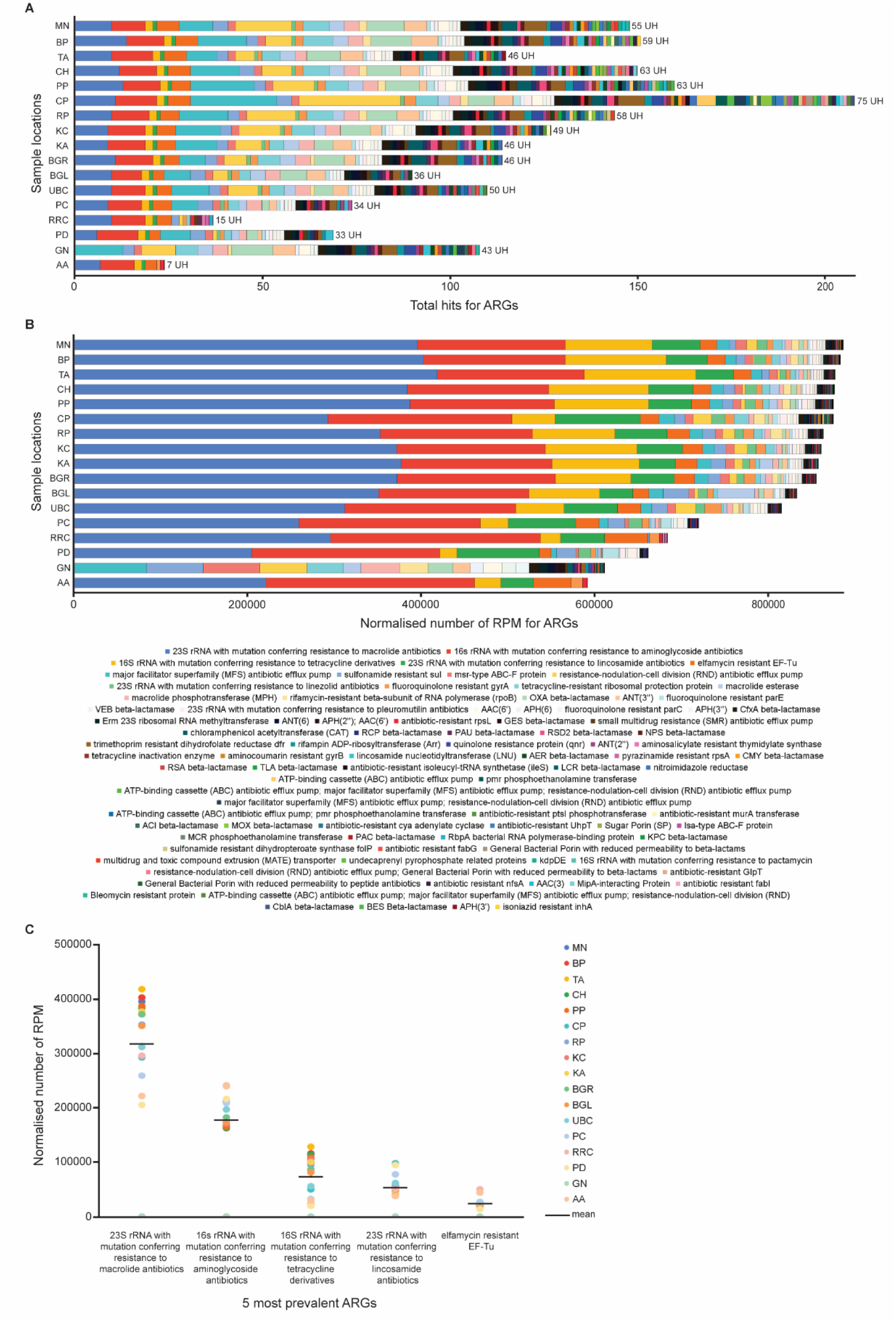
Analysis of antimicrobial resistance genes (ARGs) across the 17 locations in the city: **A)** Bar diagram showing the distribution pattern of the ARG hits in the 17 locations. UH= Unique Hits. **B)** Bar diagram showing the distribution pattern of the prevalence of the ARGs in the 17 locations. Y axis represents the normalised reads per million of the ARGs. **C)** Scatter plot showing the distribution of normalized reads per million of the 5 most prevalent ARGs across the 17 locations.

Based on the normalized reads count, the most prevalent ARGs across all locations, except GN, was ‘23S rRNA with mutation conferring resistance to macrolide antibiotics’ with a prevalence of 39.7% and mean RPM of 318105.3, followed by ‘16s rRNA with mutation conferring resistance to aminoglycoside antibiotics’ with a prevalence of 22.2% and mean RPM of 177841.1, ‘16S rRNA with mutation conferring resistance to tetracycline derivatives’ with a prevalence of 9.2% and mean RPM of 73361.6, ‘23S rRNA with mutation conferring resistance to lincosamide antibiotics’ with a prevalence of 6.7% and mean RPM of 53592.2 and ‘elfamycin resistant EF-Tu’ with a prevalence of 2.9 % and mean RPM of 23500.7 (Figure 2B, C, Supplementary Figure 3, Supplementary table 5). It is worth noting that 80.65% of the ARG reads belonged to these 5 ARGs across all samples, and except for one location-GN, all other locations had more than 75% of the normalized reads annotated to these top 5 ARGs (Supplementary Figure 3).

### Macrolide antibiotics have developed the highest antimicrobial resistance

72 unique drug classes that developed resistance in the pathogens were seen across the 17 locations in the city (Supplementary Table 1 and 6). The location CP showed the highest diversity of antimicrobial resistant drug classes with a total of 208 hits, of which 60 were unique, while the location AA showed the lowest diversity of antimicrobial resistant drug classes with only 7 unique hits out of the 24 hits observed (Figure 3A, Supplementary Figure 4, Supplementary Table 1 and 6). These unique hits were for aminoglycoside antibiotic, macrolide antibiotic, tetracycline antibiotic, fluoroquinolone antibiotic, elfamycin antibiotic, lincosamide antibiotic and macrolide; lincosamide antibiotic together. Except for elfamycin antibiotic and macrolide; lincosamide antibiotic, which were not seen in the location GN, the other 5 drug class resistance was observed in all the other 16 locations across the city. The normalized read counts across the locations suggested that macrolide antibiotic has developed the highest resistance across the city, with a mean RPM of 321214.1 constituting to 40.1% of the drug class reads, followed by aminoglycoside antibiotic with a mean RPM of 195416.3 constituting to 24.4% of the drug class reads, tetracycline antibiotic with a mean RPM of 90148.7 constituting to 11.3% of the drug class reads, lincosamide antibiotic with a mean RPM of 53873.1 constituting to 6.7% of the drug class reads and elfamycin antibiotic with a mean RPM of 23500.7 constituting to 2.9% of the drug class reads (Figure 3 B, C, Supplementary Figure 5 and Supplementary Table 7). Except for elfamycin antibiotic resistance, which was not seen in the location GN, the other four antimicrobial resistant drug classes were seen in all the locations. It is worth noting that 85.4% of the reads belonged to these top 5 drug classes for all the samples taken together and 64.5% reads belonged to macrolide and aminoglycoside antibiotics taken together for all the locations.

**Figure 3:**
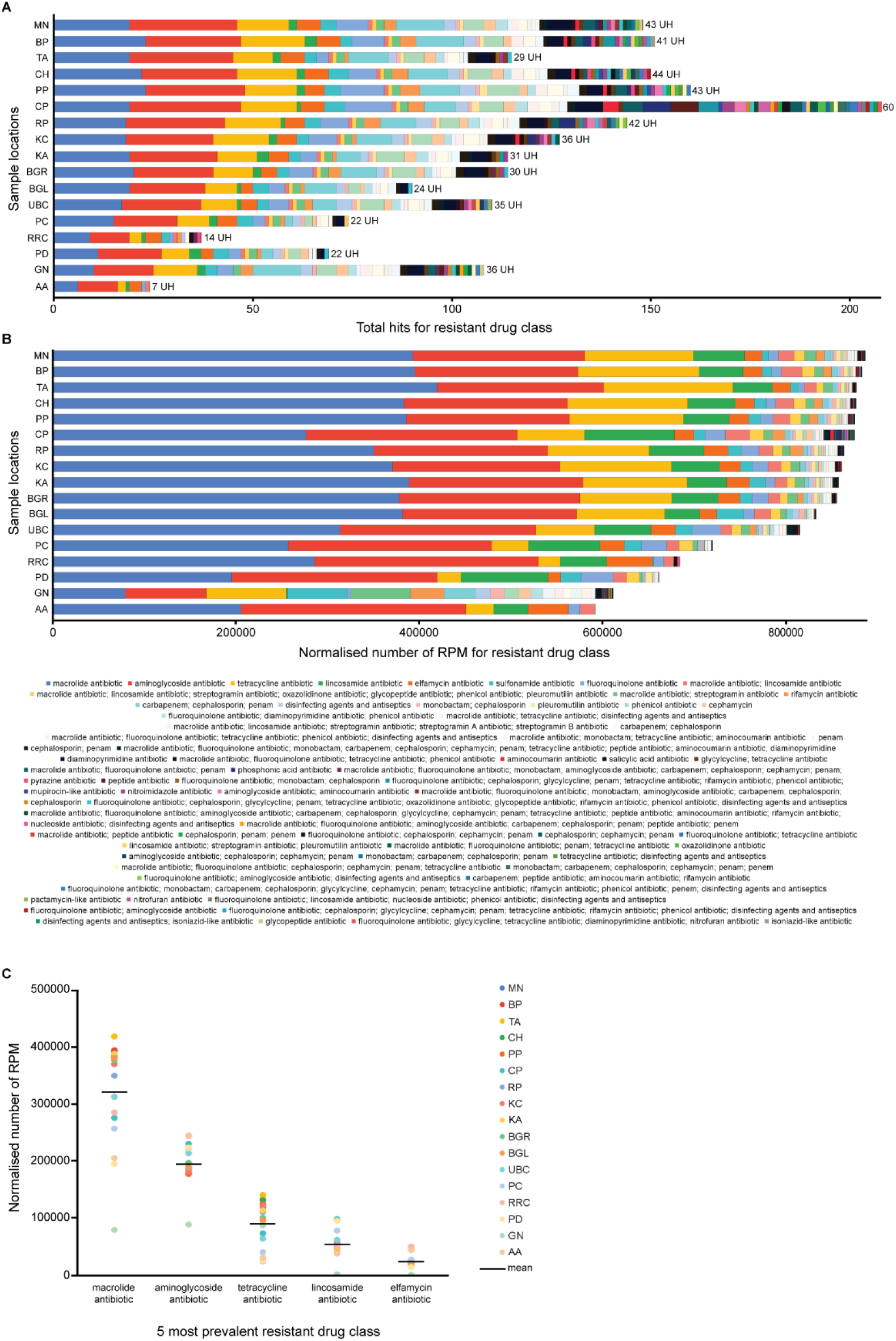
Analysis of antimicrobial resistant drug classes across the 17 locations: **A)** Bar diagram showing the distribution pattern of the hits of antimicrobial resistant drug classes in the 17 locations. UH= Unique Hits. **B)** Bar diagram showing the distribution pattern of the prevalence of the antimicrobial resistant drug classes in the 17 locations. Y axis represents the normalised reads per million of the antimicrobial resistant drug classes. **C)** Scatter plot showing the distribution of normalized reads per million of the 5 most prevalent antimicrobial resistant drug classes across the 17 locations.

### Analysis of pathogens carrying ARGs

Based on the output of the CARD, we analysed the pathogens carrying the ARGs in wastewater. 89 unique pathogens carrying the ARGs were observed in wastewater samples across the city (Supplementary Table1 and 8). The location PP that had 161 hits, showed the highest number of unique hits of 73 for pathogens amongst the 17 locations, followed by 66 unique pathogens hits in each in the location CP (out of 208 hits), BP (out of 151 hits) and CH (out of 150 hits). The location AA showed the least number of unique hits of 22 out of 24 for pathogens carrying ARGs (Figure 4A, Supplementary Figure 6, Supplementary Table 1 and 8). The pathogens *Escherichia coli, Salmonella enterica* and *Campylobacter jejuni* were seen in all the 17 locations, while *Pseudomonas aeruginosa, Neisseria gonorrhoeae, Vibrio cholerae, Helicobacter pylori, Clostridioides difficile, Enterococcus faecium, Burkholderia dolosa, Chlamydia psittaci, Chlamydia trachomatis, Chlamydomonas reinhardtii, Cutibacterium acnes, Moraxella catarrhalis, Neisseria meningitidis, Pasteurella multocida, Streptomyces cinnamoneus* and *Vibrio fluvialis* were seen in 16 out of the 17 locations (Figures 4 A, Supplementary Figure 6 and Supplementary Table 8). Based on the CARD output, the pathogens that had the highest normalized read count across the 17 locations was *Helicobacter pylori* with a mean RPM of 114041.7 constituting 14.2% of the pathogens reads, followed by *Campylobacter jejuni* with a mean RPM of 95689.3 constituting 11.9% of the pathogens reads, *Escherichia coli* with a mean RPM of 89183.8 constituting 11.1% of the pathogens reads, *Neisseria gonorrhoeae* with a mean RPM of 84860.7 constituting 10.6% of the pathogens reads and *Neisseria meningitidis* with a mean RPM of 57879.5 constituting 7.2% of the pathogens reads (Figure 4 B, C, Supplementary Figure 7, Supplementary Table 8). Except for the location GN that did not show *Helicobacter pylori, Neisseria gonorrhoeae* and *Neisseria meningitidis*, all the other 16 locations showed all these top 5 pathogens, which in total constituted to 55.1% of the pathogens reads.

**Figure 4:**
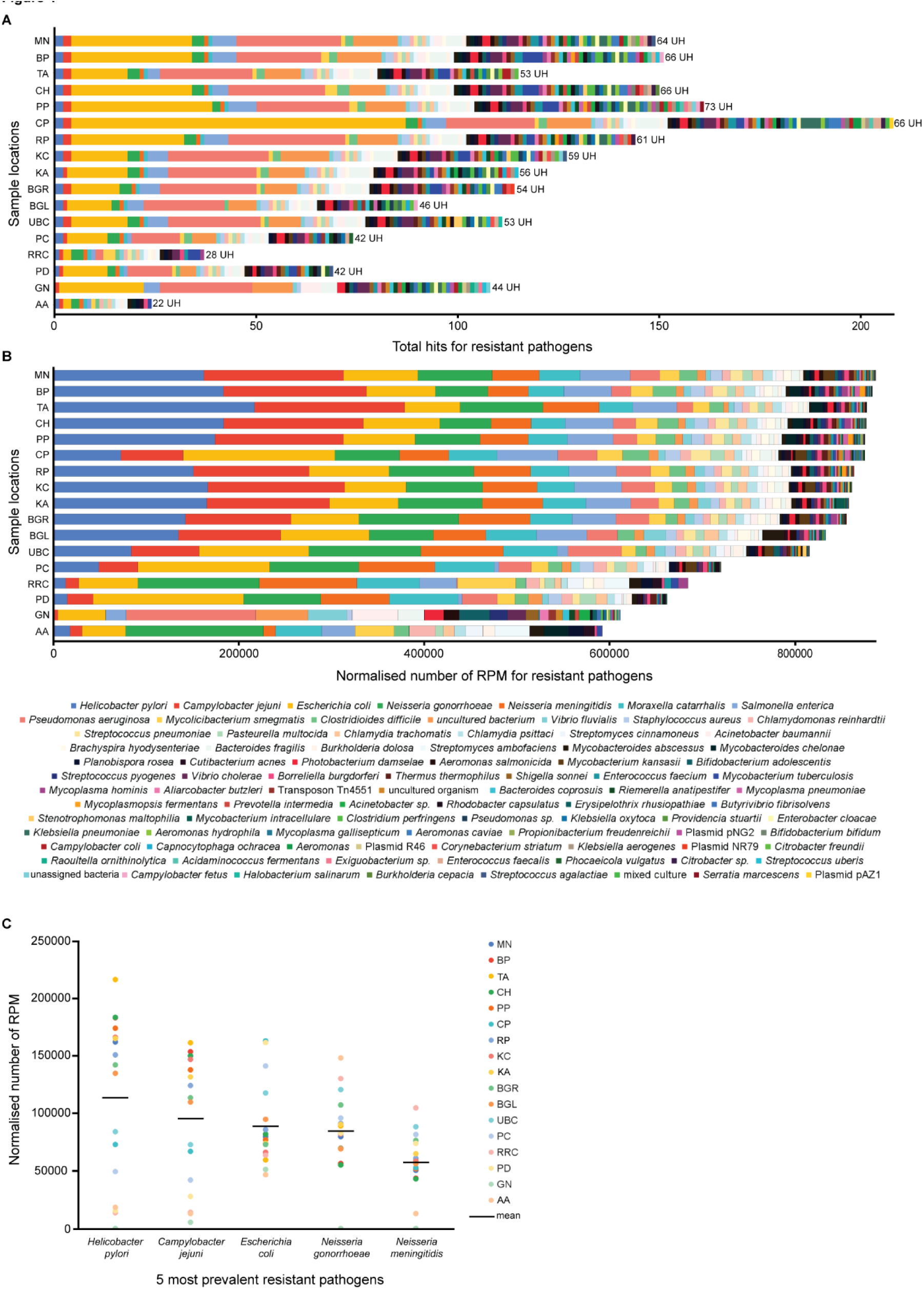
Analysis of pathogens across the locations: **A)** Bar diagram showing the distribution pattern of the hits of pathogens in the 17 locations. UH= Unique Hits. **B)** Bar diagram showing the distribution pattern of the prevalence of the pathogens in the 17 locations. Y axis represents the normalised reads per million of the pathogens. **C)** Scatter plot showing the distribution of normalized reads per million of the 5 most prevalent pathogens across the 17 locations.

### Antibiotic target alteration is the main mechanism for gaining antimicrobial resistance in the pathogens

We analysed the mechanisms through which the pathogens developed antimicrobial resistance, in wastewater samples across the city. 13 unique resistance mechanisms were observed in the 17 samples across the city (Supplementary Table 1 and 10). The location CP showed the maximum number of unique hits of 12 out of the total 208 hits for resistance mechanisms, while AA showed only one unique hit (out of 24 hits) of resistance mechanism which was antibiotic target alteration, common in all the 17 locations (Figure 5 A, Supplementary Figure 8, Supplementary Table 10). The resistance mechanism with the highest read count across all locations was antibiotic target alteration with a mean RPM of 679418 constituting 84.8% of the resistance mechanism reads across all the locations taken together (Figure 5 B, C, Supplementary Figure 9 and Supplementary Table 11). Individually, all locations, except for GN had more than 82% of the reads mapped to antibiotic target alteration mechanism. Apart from antibiotic target alteration, other AMR mechanism like antibiotic inactivation, antibiotic efflux, antibiotic target replacement accounted for 6.3%, 3.4%, 2.4%, and 2.1%, respectively, of reads associated with resistance mechanism. Despite much interest of antibiotic efflux pumps in antibiotic resistance mechanism, our data suggests that antibiotic target alteration via mutations seems to be the most probable pathway for AMR amongst pathogens ^27, 28^.

**Figure 5:**
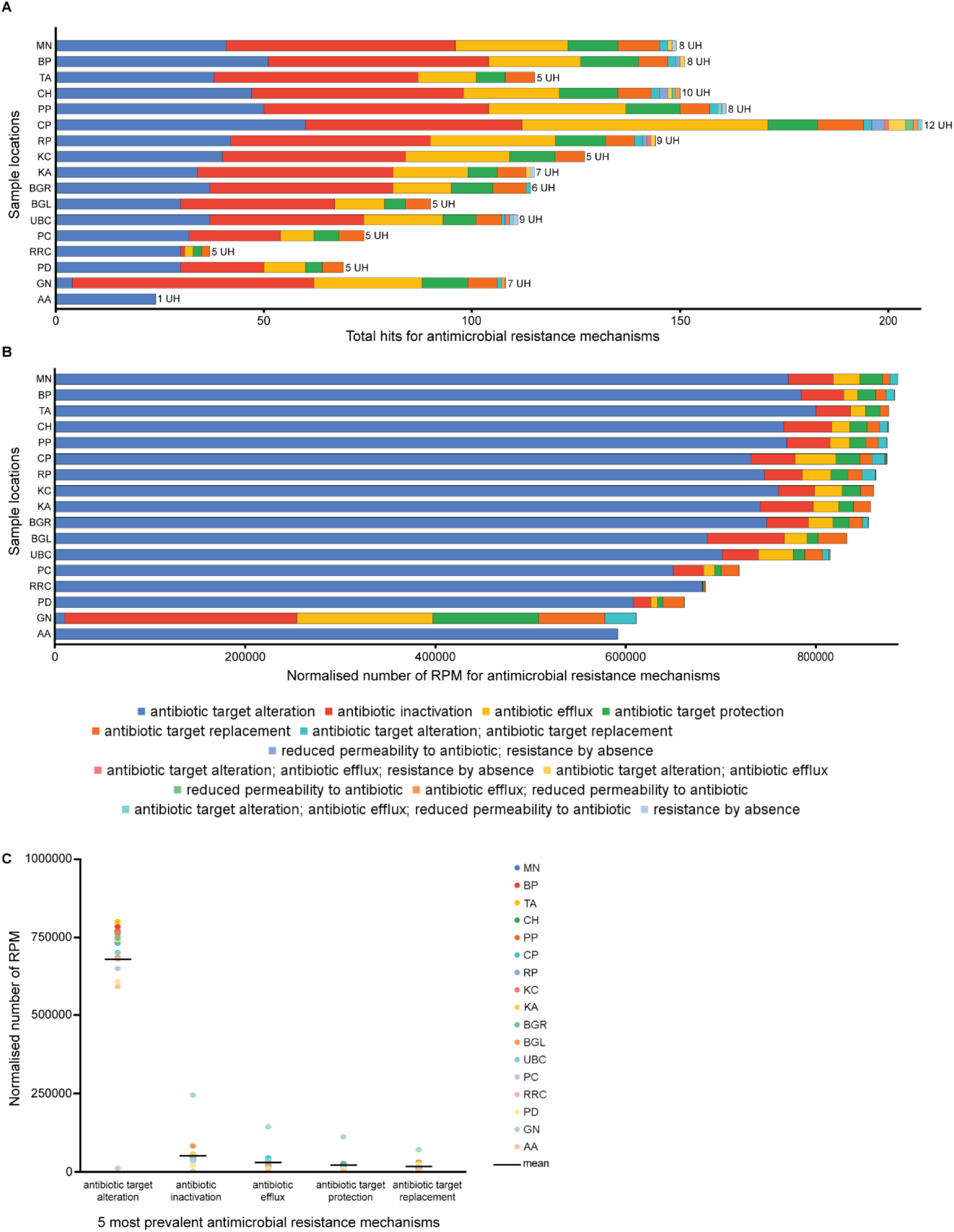
Analysis of antimicrobial resistance mechanisms across the locations: **A)** Bar diagram showing the distribution pattern of the hits of antimicrobial resistance mechanisms in the 17 locations. UH= Unique Hits. **B)** Bar diagram showing the distribution pattern of the prevalence of the antimicrobial resistance mechanisms. Y axis represents the normalised reads per million of the antimicrobial resistance mechanisms. **C)** Scatter plot showing the distribution of normalized reads per million of the 5 most prevalent antimicrobial resistance mechanisms across the 17 locations.

### Principal component analysis of the AROs, ARGs, drug classes, pathogens and resistance mechanisms

In order to understand the distribution patterns of AROs, ARGs, drug classes, pathogens and resistance mechanisms at city level, we performed principal component analysis (PCA) of the data points. We observed that the data variation across the locations GN and CP was maximum, and could explain 57.22 %, 64.1%, 67.54%, 75.96% and 55.84% variations in the data for AROs, ARGs, drug classes, resistance mechanisms and pathogens respectively. Accordingly, all locations in the city, except for GN and CP, clustered together with respect to the AROs, ARGs, drug classes and resistance mechanisms indicating that except for the two locations, the city in general showed an even distribution pattern. As expected, the distribution of the pathogens was not tightly clustered across the locations in the city (Figure 6A-E and Supplementary figure 10), suggesting a diverse pathogen profile at different locations.

**Figure 6:**
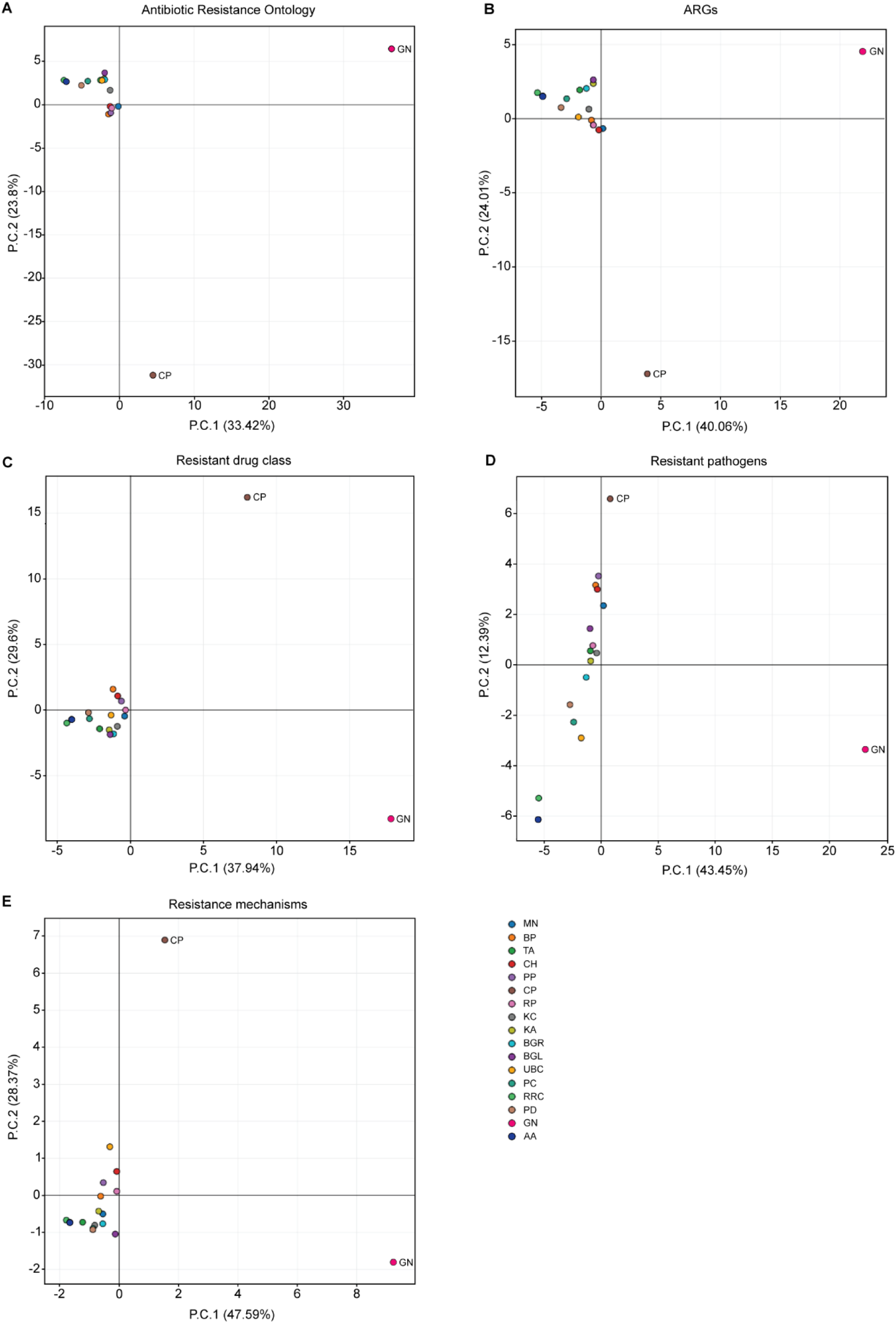
Principal component analysis (PCA) of the A) AROs, B) ARGs, C) antimicrobial resistant drug classes, D) pathogens and E) antimicrobial resistance mechanisms across the 17 locations. PC1 and PC2, that explain the maximum variation amongst the locations, are shown on the X and Y axes respectively.

### Assessment of global and Indian AMR critical pathogens

We next analysed the prevalence of the AMR critical pathogens and the corresponding drug classes for which resistance was observed our samples. The World Health Organization (WHO) has published a list of antibiotic-resistant “priority pathogens”^29^. This list is divided into 3 categories based on the severity-critical, high and medium priority pathogens. The critical priority pathogens are *Acinetobacter baumannii*, *Pseudomonas aeruginosa* and members of the family *Enterobacteriaceae*, that have developed resistance against multiple antibiotics, especially carbapenem, and are of specific concerns in settings like hospitals and nursing homes that house patients requiring devices like ventilators and blood catheters. These are also associated with nosocomial infections and are resistant to even the third generation cephalosporins. The high priority pathogens include vancomycin resistant *Enterococcus faecium*, methicillin and vancomycin resistant *Staphylococcus aureus*, clarithromycin resistant *Helicobacter pylori*, fluoroquinolone resistant *Campylobacter*, fluoroquinolone resistant *Salmonellae*, and cephalosporin and fluoroquinolone resistant *Neisseria gonorrhoeae*. The medium priority pathogens include penicillin-non-susceptible *Streptococcus pneumoniae,* ampicillin resistant *Haemophilus influenzae* and fluoroquinolone resistant *Shigella*. The India priority pathogen list developed by WHO Country Office for India in collaboration with Department of Biotechnology, Government of India, also includes *Neisseria meningitidis* as a pathogen of medium priority along with the above mentioned pathogens of concern^30^.

We analysed the prevalence of these pathogens of concern in our 17 wastewater samples. All of these pathogens except for *Haemophilus influenzae* were detected in wastewater in the city (Figure 7). The first critical priority *Enterobacteriaceae* family member *Escherichia coli* alone showed resistance to 38 drug classes in our samples. The most prevalent resistant drug classes were lincosamide with 92599 RPM, followed by aminoglycoside with 17157.7 RPM, elfamycin with 12983.2 RPM, macrolide with 12814.6 RPM, rifamycin with 5229.6 RPM and carbapenem; cephalosporin; penam with 1635.9 RPM. *Klebsiella pneumoniae,* another member of the *Enterobacteriaceae* family showed resistance to 9 drug classes in our samples. The top 5 classes were aminoglycoside antibiotic with 185.2 RPM, monobactam; carbapenem; cephalosporin; penam with 97.9 RPM, carbapenem; cephalosporin; penam with 96.6 RPM, carbapenem; peptide antibiotic; aminocoumarin antibiotic; rifamycin antibiotic with 58.6 RPM and macrolide antibiotic; peptide antibiotic with 51.3 RPM. Another *Enterobacteriaceae* family member *Enterobacter cloacae* showed resistance to phenicol antibiotic with 412.9 RPM. The critical priority pathogen *Pseudomonas aeruginosa* showed resistance to 21 antibiotic agents in our samples. The most prevalent was fluoroquinolone antibiotic with RPM of 15433.2, followed by aminoglycoside antibiotic with 10096.3 RPM, monobactam; cephalosporin with 5524.8 RPM, disinfecting agents and antiseptics with 4105.9 RPM, fluoroquinolone antibiotic; diaminopyrimidine antibiotic; phenicol antibiotic with 2930.2 RPM and carbapenem; cephalosporin; penam with 2671.8. The critical priority pathogen *Acinetobacter baumannii* showed resistance to 4 drug classes-macrolide antibiotic; streptogramin antibiotic with RPM of 9753.2, followed by carbapenem; cephalosporin; penam with 141.2 RPM, aminoglycoside antibiotic with 48.6 RPM and glycopeptide antibiotic with 5.3 RPM. Importantly, except for *Enterobacter cloacae,* all other critical pathogens showed resistance to carbapenem and cephalosporin along with multiple other drug classes in our samples.

**Figure 7:**
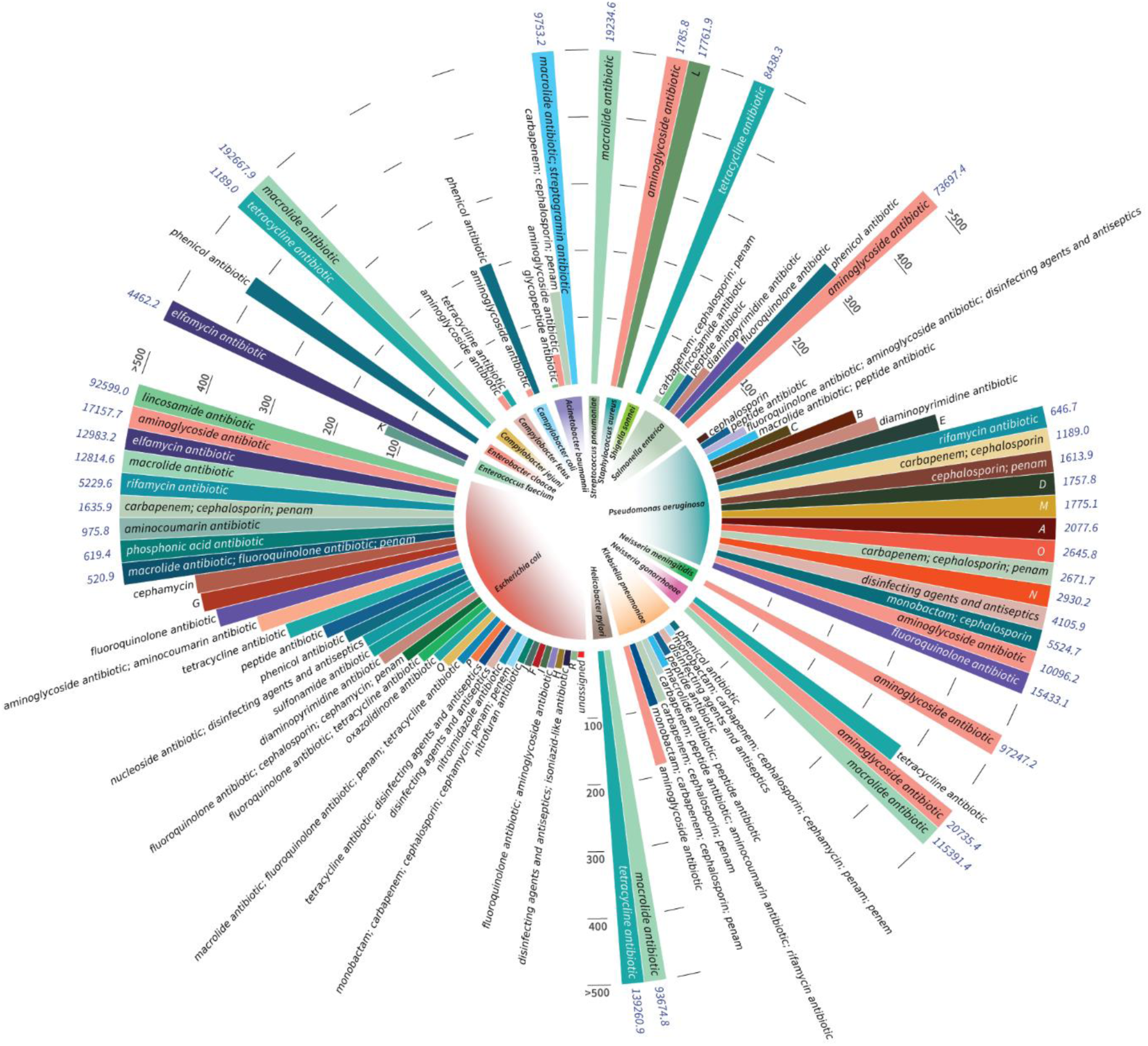
Plot showing the abundance of the WHO Global and Indian Priority Pathogens and the respective resistant drug classes seen in the samples

The high priority pathogen *Enterococcus faecium* showed resistance to elfamycin antibiotic with RPM of 4462.3 and macrolide antibiotic; lincosamide antibiotic; streptogramin antibiotic; streptogramin A antibiotic; streptogramin B antibiotic with RPM of 129.2 in our samples. The pathogen *Staphylococcus aureus* showed resistance to aminoglycoside antibiotic with 1785.9 RPM and macrolide antibiotic; lincosamide antibiotic; streptogramin antibiotic; oxazolidinone antibiotic; glycopeptide antibiotic; phenicol antibiotic; pleuromutilin antibiotic with 17761.9 RPM. *Helicobacter pylori* showed resistance to tetracycline antibiotic with 139261 RPM and macrolide antibiotic with 93674.9 RPM. We observed three *Campylobacter* species in our samples-*Campylobacter jejuni* resistant to macrolide antibiotic with 192668 RPM and tetracycline antibiotic with 1189 RPM, *Campylobacter coli* resistant to phenicol antibiotic with 207.8 RPM and aminoglycoside antibiotic with 11.3 RPM, *Campylobacter fetus* resistant to tetracycline antibiotic with 26.6 RPM and aminoglycoside antibiotic with 24 RPM. The high priority pathogen *Salmonella enterica* showed resistance to 7 drug classes-aminoglycoside antibiotic with 73697.5 RPM, phenicol antibiotic with 320.4 RPM, fluoroquinolone antibiotic with 146.6 RPM, diaminopyrimidine antibiotic with 84.6 RPM, peptide antibiotic with 62 RPM, lincosamide antibiotic with 57.3 RPM and carbapenem; cephalosporin; penam with 11.3 RPM. The pathogen *Neisseria gonorrhoeae* showed resistance to three drug classes in our samples-macrolide antibiotic with 115391.5 RPM, aminoglycoside antibiotic with 20735.4 RPM and tetracycline antibiotic with 380.4 RPM.

Two medium priority pathogens from the WHO list were observed in our samples-*Streptococcus pneumoniae* resistant to macrolide antibiotic with 19234.7 RPM and *Shigella sonnei* resistant to tetracycline antibiotic with 8438.3 RPM were observed in our samples. Along with these, the medium pathogen from the WHO-Indian list, *Neisseria meningitidis* resistant to aminoglycoside antibiotic with 97247.2 RPM, was also seen in the samples.

## Discussion

AMR is a global concern, and its geographical surveillance is extremely important in order to design appropriate mitigation strategies. In this study, we have comprehensively elucidated the AMR landscape of an Indian metropolitan city using the culture-independent metagenomics approach. We chose the sample collection from open drainage sites over sewage treatment plants (STP) for the study, as we wanted to assess and set up a surveillance system that works for cities, towns and villages in India, that do not have a well-planned sewerage system with STPs. Open drainage system is more common in semi-urban and rural settings in the country, and it receives wastes not only from humans, but also from animals, industries, poultries and farming practices. Thus, samples from open drainage sites provide a comprehensive overview of the environment and the associated microbiome carrying the antimicrobial resistance. Previous metagenomics-based studies that assessed the global landscape of AMR by wastewater surveillance observed a high degree of local and regional resistomes over 3 years ^31, 32^. They observed high prevalence of resistance to macrolides, followed by aminoglycosides, tetracycline and B-lactams. Interestingly, our samples show a similar pattern of resistance, re-emphasizing that there is a need to develop newer strategies to combat the problem of these resistant drugs. A recent bacterial culture based study that assessed the antimicrobial resistance pattern in wastewater in 4 Indian cities also observed substantial resistance against these drug classes ^33^. In our study, we observed that the antimicrobial resistance patterns are broadly similar over the geographical locations in the city. A co-relation analysis of the number of hospitals in 2 km radius of our sampling locations with the number of AROs suggests that there is a positive co-relation (Pearsons co-efficient of co-relation: 0.3) (Figure 8, Supplementary Table 13). The co-relation is stronger (Pearsons co-efficient of co-relation: 0.8) when the two sites PD and RRC are removed from the analysis.

**Figure 8:**
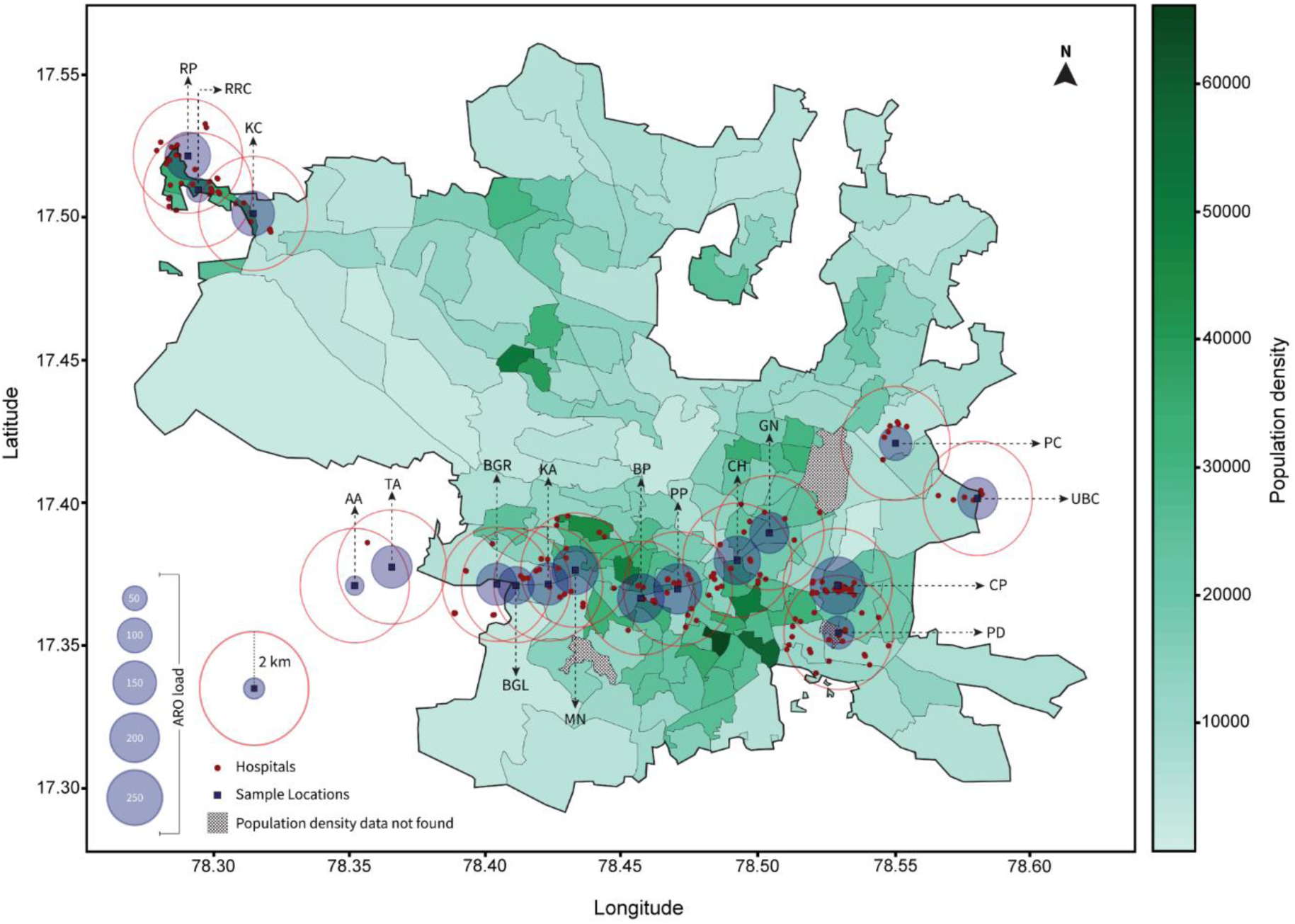
Ward-wise population density map of the city showing the antimicrobial resistance ontology (ARO) load at the 17 sampling locations. The blue squares indicate the geographical co-ordinates of the 17 wastewater sampling sites. The translucent blue circles around the sites indicate the ARO loads at the respective sites. The red dots indicate the geographical co-ordinates of the hospitals in the 2 kilometres radius of the sites. The red lined circles indicate the 2 kilometres radius around the sampling sites.

Network graph of our data of ARGs, drug classes, pathogens and resistance mechanisms revealed that ARGs have wide ranges of host pathogens (Figure 9, Supplementary Table 14). The most prevalent ARG in our samples ‘23S rRNA with mutation conferring resistance to macrolide antibiotics’ was present in multiple pathogens including the AMR critical pathogens *Campylobacter jejuni, Neisseria gonorrhoeae, Helicobacter pylori* and *Streptococcus pneumoniae*. The next prevalent ARG, ‘16s rRNA with mutation conferring resistance to aminoglycoside antibiotics’ was present in the AMR critical pathogens *Escherichia coli, Neisseria gonorrhoeae, Neisseria meningitidis, Salmonella enterica* and, also in *Mycobacterium tuberculosis* amongst others. Similarly, single pathogens harbored a wide range of ARGs and were resistant to multiple drug classes. For example, *Escherichia coli* alone harbored 43 ARGs and was resistant to 38 drug classes, *Pseudomonas aeruginosa* harbored 21 ARGs and was resistant to 21 drug classes, *Klebsiella pneumoniae* had 9 ARGs and was resistant to 9 drug classes, and *Salmonella enterica* harbored 9 ARGs and was resistant to 7 drug classes in our samples. The ESKAPE pathogens (*Enterococcus faecium, Staphylococcus aureus, Klebsiella pneumoniae, Acinetobacter baumannii, Pseudomonas aeruginosa,* and *Enterobacter spp*.) are responsible for nosocomial infections worldwide and there is a growing concern of multidrug resistance in these pathogens ^34, 35^. *Streptococcus pneumoniae* infections along with ESKAPE pathogens were associated with more than 3 million AMR related deaths globally in 2019 ^19^. In our data, we found that *Streptococcus pneumoniae* and ESKAPE pathogens accounted for 8.2% of the normalized pathogens reads. Individually, the highest number of reads were for *Pseudomonas aeruginosa* (4.4%), followed by *Staphylococcus aureus* (1.3%), *Streptococcus pneumoniae* (1.2%), *Acinetobacter baumannii* (0.9%), *Enterococcus faecium* (0.3%), *Enterobacter cloacae* (0.035%), and *Klebsiella pneumoniae* (0.035%). It is known that extrapulmonary manifestations of tuberculosis can lead to the discharge of *Mycobacterium tuberculosis* in the wastewater ^36^. We too observed *Mycobacterium tuberculosis* in our wastewater samples, with around 0.27% of normalized pathogens reads. The pathogen was found in all locations except GN and BGL. Our samples also revealed that a significant proportion of normalized pathogen reads, 26.1%, belonged to *Helicobacter pylori* and *Campylobacter jejuni*, the two pathogens often associated with zoonosis. Along with humans, *Helicobacter spp* and *Campylobacter spp* are also known to infect non-human hosts like livestock, rodents and domestic animals, and thus have an important role to play in disease spillovers ^37, 38^. Taken together, our data demonstrates that many major pathogens are evolving or acquiring antibiotic resistance genes to evade antibiotics of all major classes in multiple hosts.

**Figure 9:**
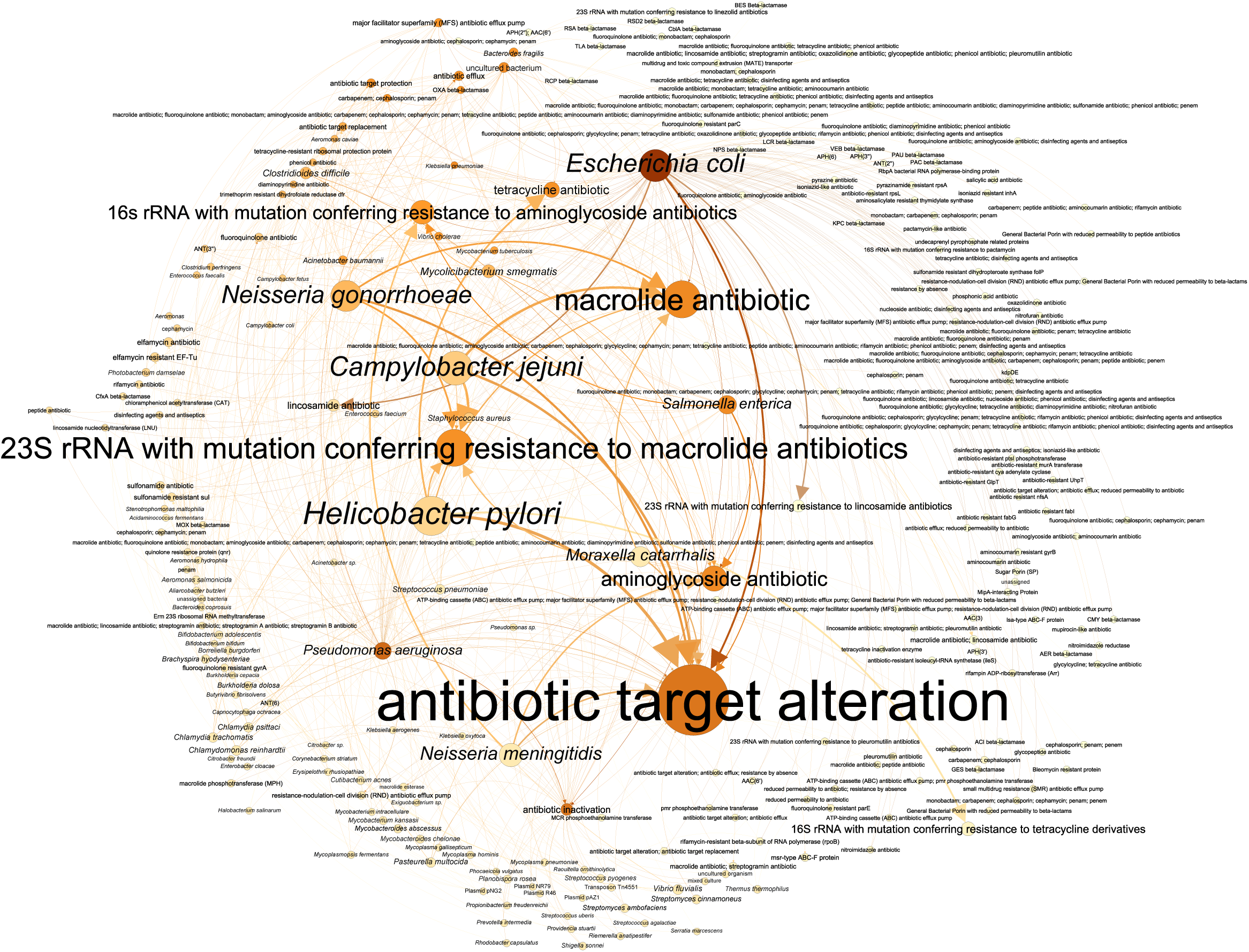
Network plot showing the ARGs, antimicrobial resistant drug classes, pathogens and antimicrobial resistance mechanisms across the city. Nodes represent the attributes-ARGs, antimicrobial resistant drug classes, pathogens and antimicrobial resistance mechanisms and edges represent the relationship between the attributes. The colour intensity of the nodes is proportional to the number of relations they have. The size of the node and the labelling font is proportional to the prevalence of the attribute. The thickness of the edges is proportional to the strength of the relationship between the two nodes.

The analysis of ARGs in our samples revealed that 79.3% of normalized reads belonged to mutations in 23S or 16S rRNA gene sequences. This indicates that protein synthesis inhibitor antibiotics are at a serious risk of being inactive against many pathogens ^39, 40, 41, 42^. We also found that 2.2% of normalized reads belonged to ARG which leads to resistance to β-lactam antibiotics. Amongst these reads, we found ARGs encoding for extended spectrum β-lactamases (ESBL) like VEB β-lactamase, BES β-lactamase, GES β-lactamase and KPC β-lactamase. KPC beta-lactamases is a *Klebsiella pneumoniae* carbapenemase and confers resistance to penicillins, cephalosporins, carbapenems, and most beta-lactamase inhibitors. This ARG has been associated with many epidemics around the world ^43^. Altogether, our data indicates that ARGs for major classes of antibiotics *i.e.*, macrolides, tetracyclines, sulfonamides, beta-lactams, fluoroquinolones, aminoglycosides and rifamycins are present in the open drain samples. It may be noted that our observations are based on a snapshot study of the waste-water samples collected across the city in the year 2022. Thus, the results are representative of the AMR scenario prevalent across different samples at the time of sample collection. Temporal analysis of the AMR profile across the geographical locations would give more idea on the evolution and resistance patterns across the city.

Taken together, this study gives a comprehensive picture of the antimicrobial landscape of an Indian metropolitan city with respect to the antimicrobial resistant genes, the resistant drug classes, the pathogens carrying resistance and the mechanisms of resistance, using open drain-based wastewater surveillance.

## Materials and methods

### Sampling sites and sample collection

Wastewater samples were collected from 17 sampling sites from a metropolitan city in south India that has a population of around 10 million, as of 2023. The city generates around 1650 million gallons sewage daily, of which 772 million gallons is treated through the sewage treatment plants in the city^44^. The 17 sampling sites in this study include, 10 open drains, 4 rivers and 3 lakes. The details of the sites are mentioned in Supplementary Table 12. The wastewater samples from the sites were collected in sterile bottles at morning hours between 7am-10am in January 2022, over two days, by grab sampling method, as per the procedures described earlier ^45^. The samples were transported to the laboratory and were immediately processed further.

### Samples processing

The samples were inactivated by heating in a water-bath at 60°C for 30 minutes. 50 ml of wastewater samples were concentrated using 4 g polyethylene glycol 8000 (PEG-8000; HiMedia laboratories Pvt. Ltd., Catalogue no. MB150-500G) and 0.9 g sodium chloride (NaCl; HiMedia Laboratories Pvt. Ltd, Catalogue no. MB023-500G). Briefly, PEG and sodium chloride were dissolved in the lukewarm wastewater samples, followed by overnight incubation at 4°C. The samples were centrifuged at 10,000 rpm for 30 minutes at 4°C. The pellet was used for nucleic acid extraction using the AllPrep power Viral DNA/RNA Kit (Qiagen; Catalogue No. 28000-50).

### Next generation sequencing

The eluted nucleic acids were used for library preparation using the Illumina DNA Prep Tagmentation (M) Beads 96 Samples kit (Illumina, Catalogue no. 20015880). The samples were paired-end sequenced on the Illumina Novaseq 6000 platform. Each sample was sequenced to get on an average 34 million reads. ‘FastQC (v0.11.9)’ was used for the assessment of the sequence quality and the adapter sequences were removed by ‘Trimmomatic (v0.39)’ using the default settings^46, 47^.

### Identification of the AROs, ARGs, antibiotic resistant drug classes, pathogens and resistance mechanisms from the metagenomics reads

The metagenomics reads were analysed for detection of AROs, ARGs, antibiotic resistant drug classes, pathogens and resistance mechanisms using the Resistance Gene Identifier (RGI) v5.2.0. This application mapped short reads from each sample against reference data sourced from the Comprehensive Antibiotic Resistance Database (CARD) version 3.2.5 ^25, 26, 48^. A threshold of ≥ 95% was applied to the output files and the filtered data was used for further analysis ^10^. Before plotting the graphs, the raw reads were normalized using the formula

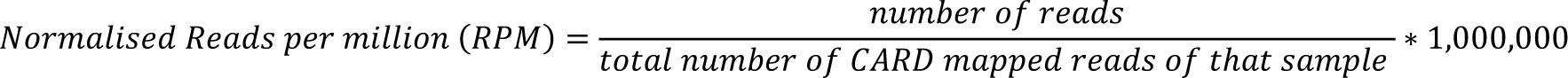

The total number of CARD mapped reads without the ≥95% coverage cut-off is used in the denominator of the above formula.

Percentage prevalence of a group was calculated by the formula

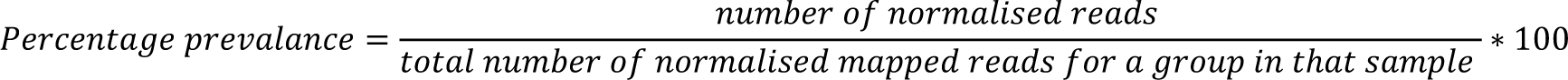

‘group’ represent ARO, ARG, drug class, pathogens or resistance mechanism in the above formula. The number of group-mapped reads with ≥95% coverage cut-off is used in the denominator of the above formula.

### Data Visualization

Data visualization was performed by excel, or Python using the Pycharm Community Edition version 2023.2.1. The libraries GeoPandas (v0.13.2), pandas (v2.0.3), NumPy (v1.24.2), matplotlib (v3.7.2), seaborn (v0.12.2), plotly (v5.16.1) were used ^10, 49, 50, 51, 52, 53, 54^. Plots were assembled and refined on Adobe Illustrator.

Network analysis was performed in Gephi (v0.10.1), using the force-directed layout algorithms Fruchterman Reingold and ForceAtlas 2 ^55^. Nodes and edges files were sorted on Python using Pandas library. Nodes represent the attributes-ARGs, antimicrobial resistant drug classes, pathogens and antimicrobial resistance mechanisms. Directed edges represent the relationship between the attributes.

### Principal component analysis

Principal component analysis (PCA) for co-variance was performed for the reads derived from CARD for AROs, ARGs, pathogens, drug class and resistance mechanisms for the 17 locations. The resulting principal components were utilized to create a two-dimensional scatter plot, providing a visual representation of the relationships and variations present in the data. PC1 and PC2, representing the first and second principal components, were selected for further analysis. K-means clustering was applied to the PCA-transformed data PC1 and PC2.

### Plotting the ward map of the city overlayed with population density

For plotting the ward-wise population density map of the city, Python programming was employed. The geographical coordinates of the respective wards were sourced from GitHub repository ^56^. The population data of the respective wards was parsed from 2011 census data obtained from the official website of Ministry of Home Affairs, Government of India ^57^. The population figures were divided by the area of the respective wards to get the ward-wise population density. The geographical co-ordinates of the wastewater sampling sites and the hospitals around the 2 km radius of the sampling sites were plotted on the ward-wise population density map using Python. The co-ordinates of the hospitals were obtained from Google Maps.

## Data Availability

All data produced in the present study are available upon reasonable request to the authors

## Acknowledgements

This study was supported by the funding from the Rockefeller Foundation, Grant Number 2021 HTH 018 and the Tata Trusts. The authors acknowledge the help of Dr. Wasimuddin, Senior Scientist, Norwegian Veterinary Institute for his technical inputs.

## Author contributions

RKM and SCM conceived the study and secured funding. NS and MKM performed the experiments. VRI performed bioinformatic analysis. MKM performed data visualization and plotting. DTS and KBT gave scientific and technical inputs. SCM and VRI drafted the initial manuscript with guidance from RKM. RKM refined the manuscript draft.

