## Supplementary Figures for "Antimicrobial resistance landscape in a metropolitan city context using open drain wastewater-based metagenomic analysis"

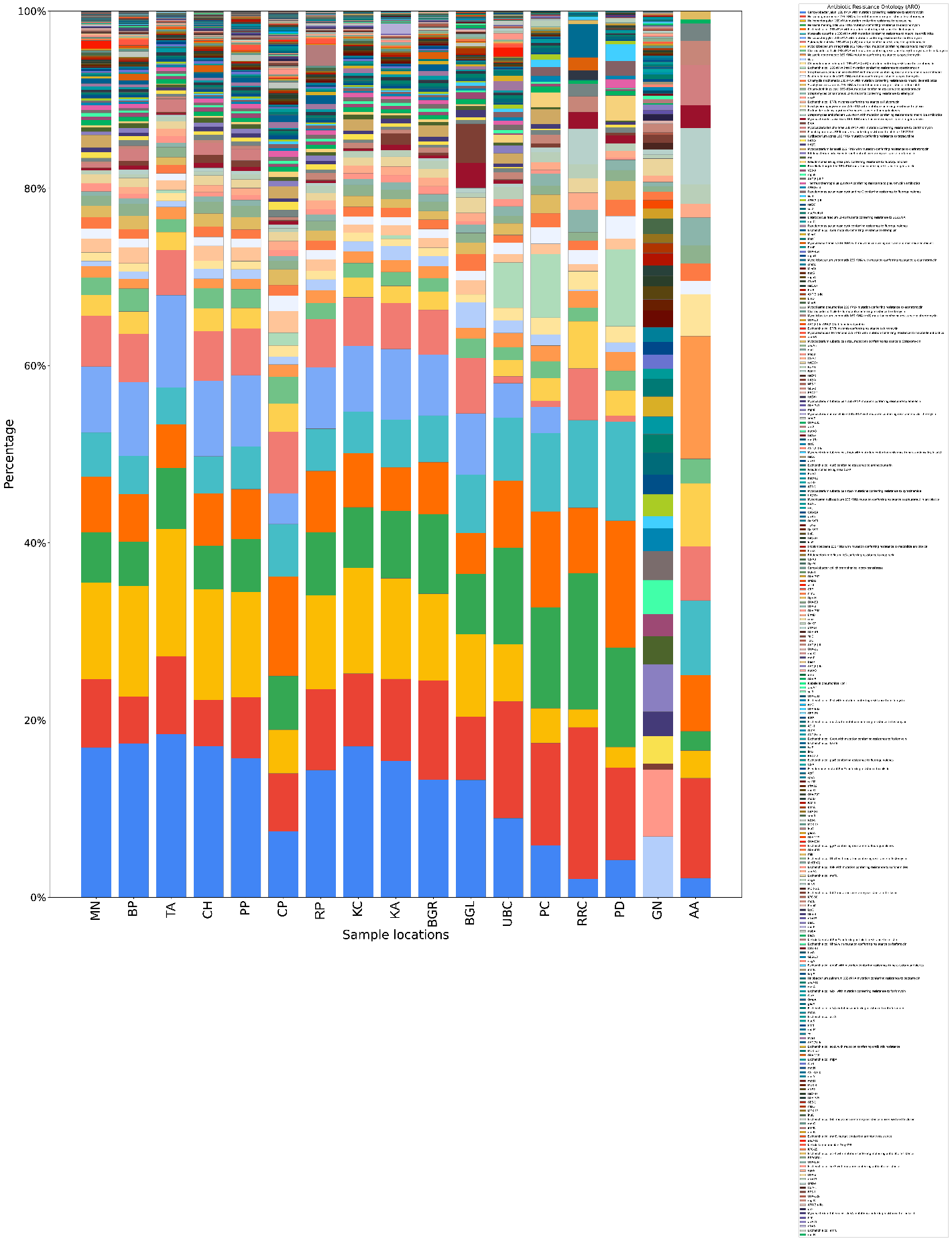

**Supplementary Figure 1:** Bar diagram showing the relative abundance of the AROs across the 17 locations based on the read counts.

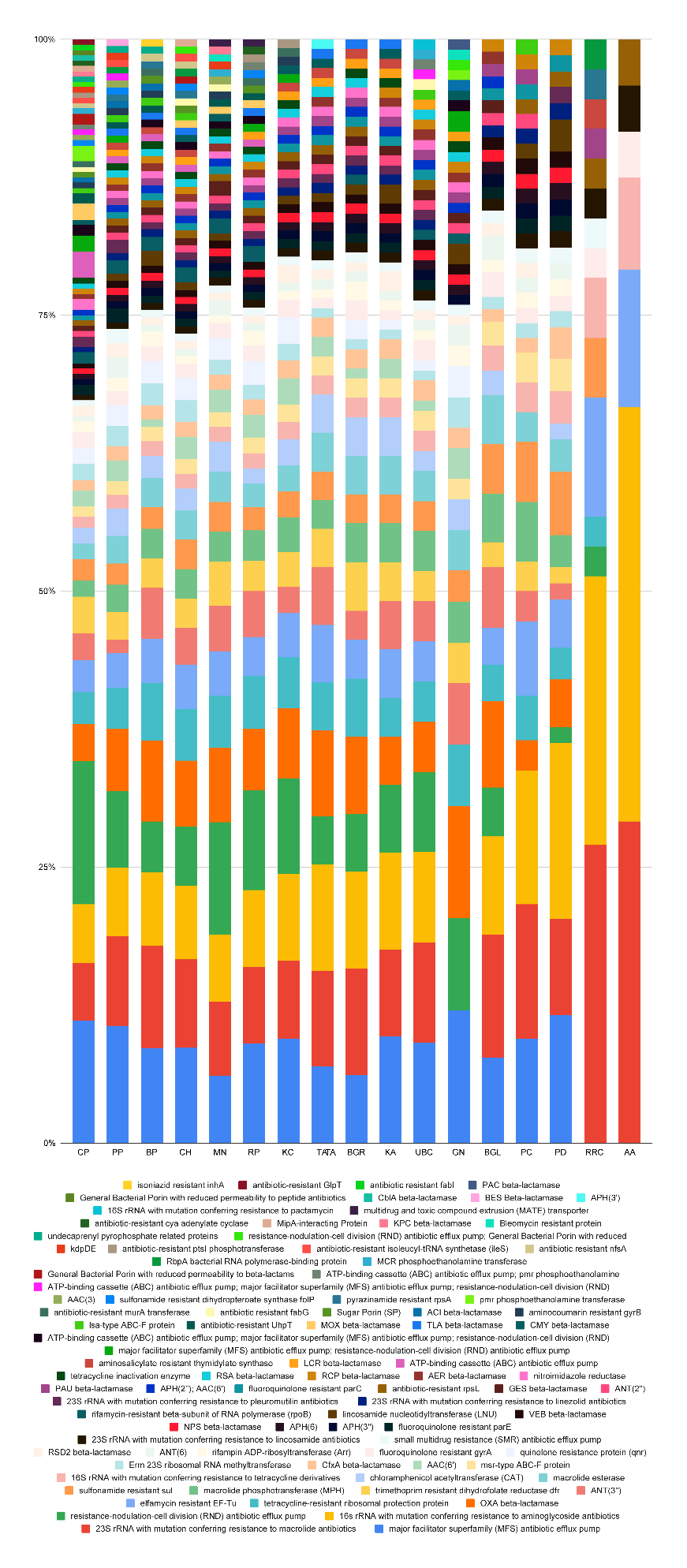

**Supplementary Figure 2:** Bar diagram showing the relative abundance of the ARGs hits across the 17 locations.

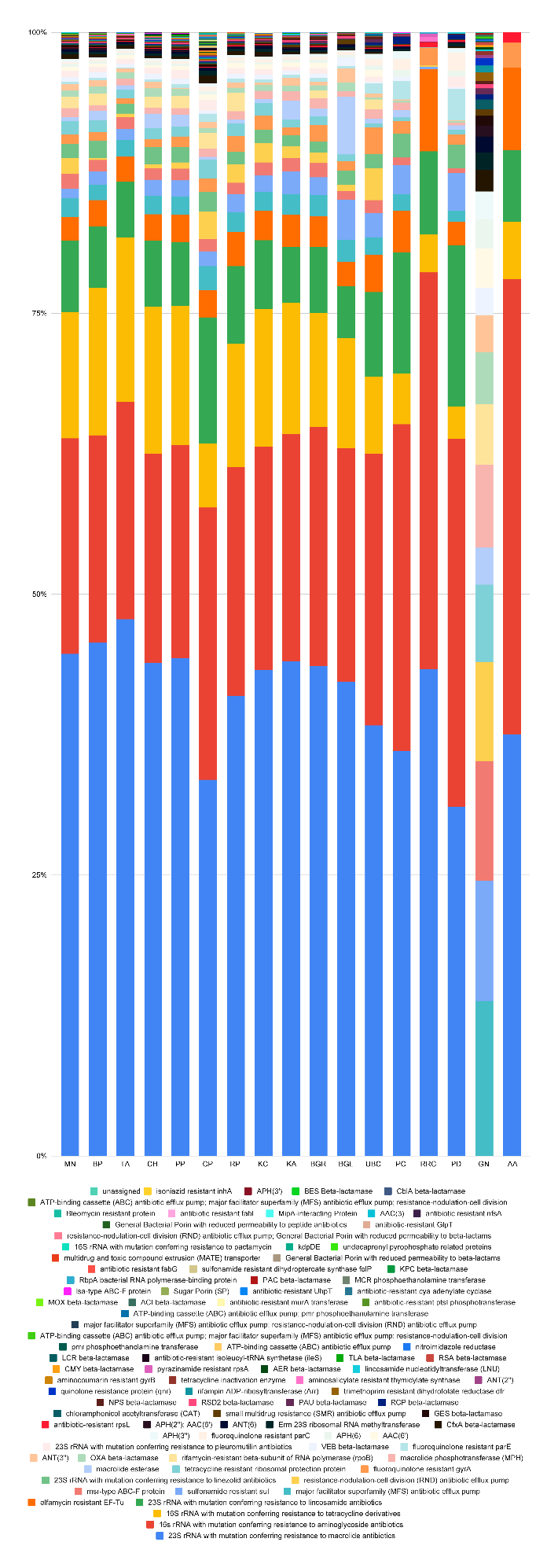

**Supplementary Figure 3:** Bar diagram showing the relative abundance of the ARGs across the 17 locations based on the read counts.

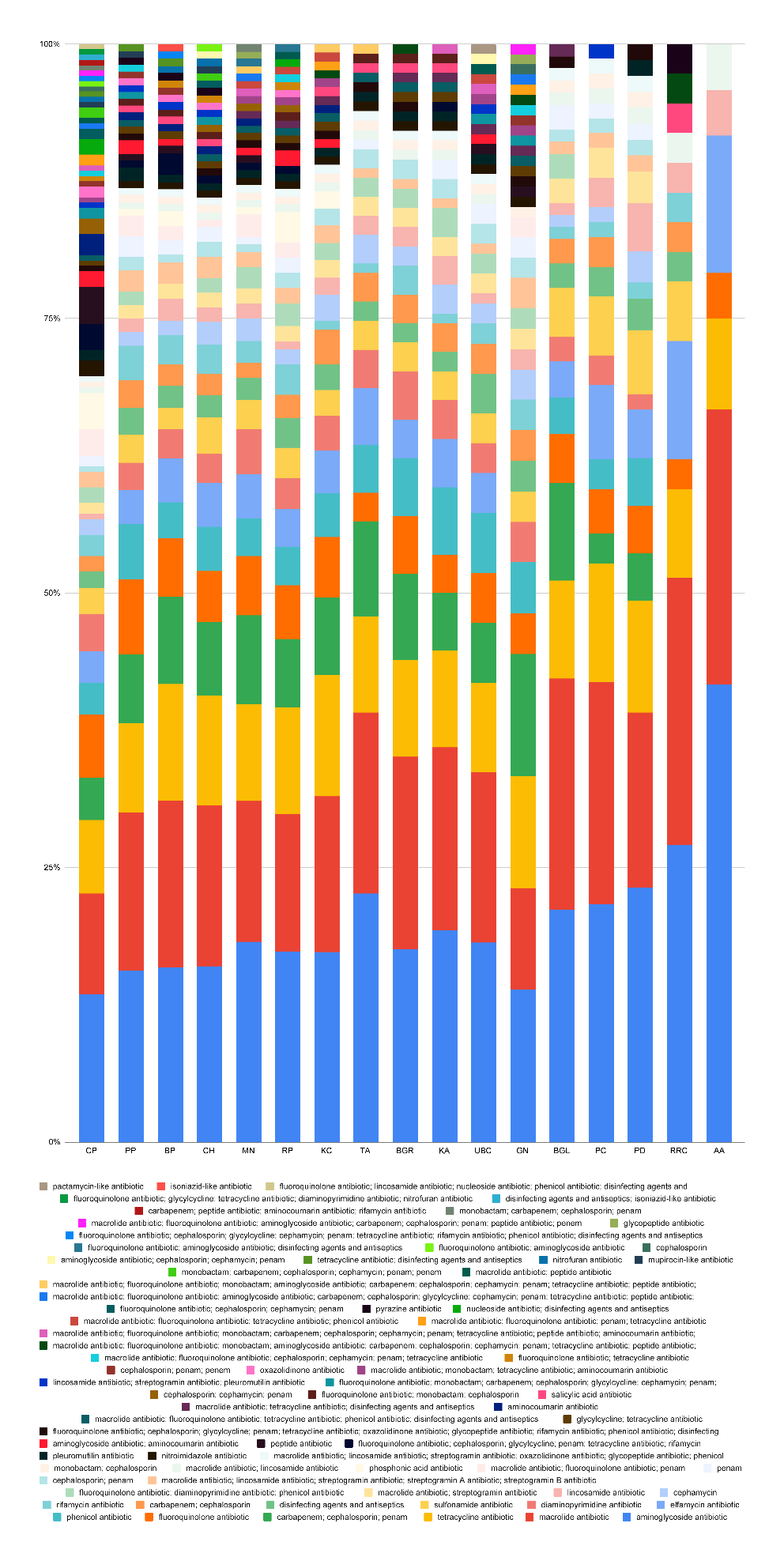

**Supplementary Figure 4:** Bar diagram showing the relative abundance of the hits of antimicrobial resistant drug classes across the 17 locations.

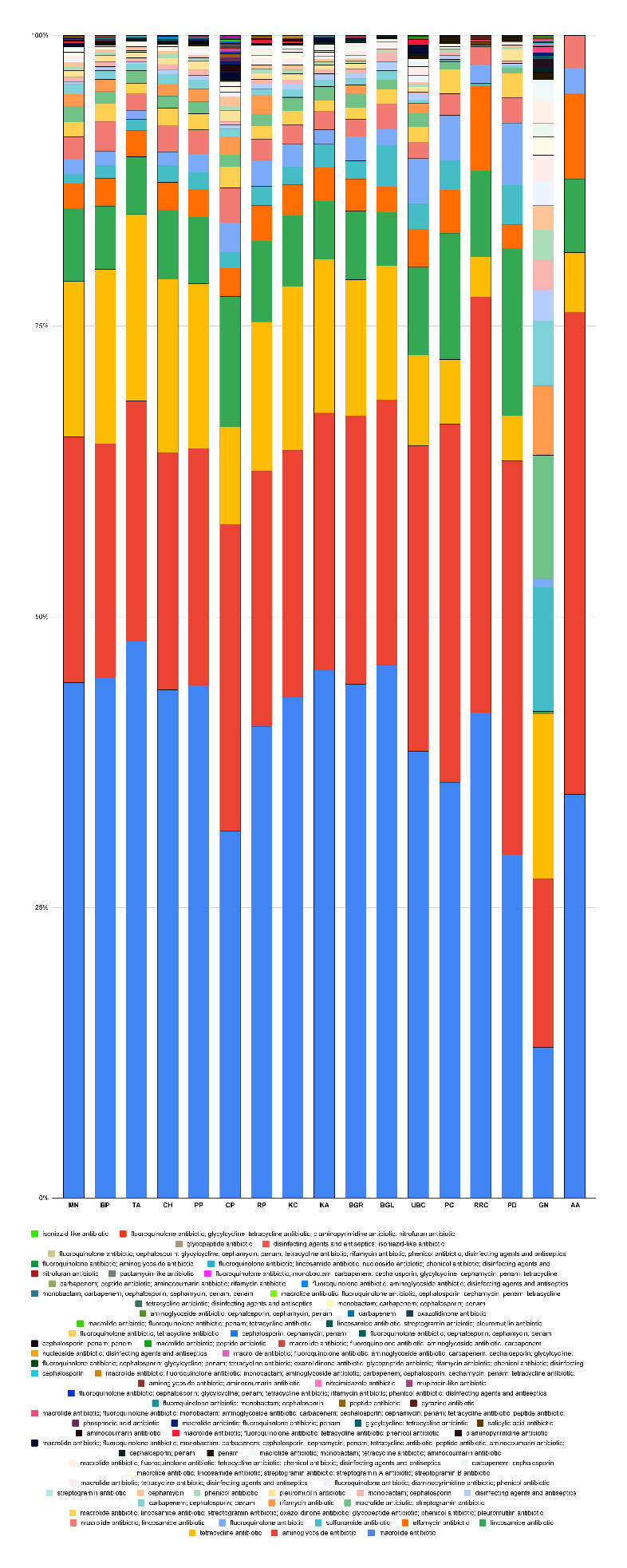

**Supplementary Figure 5:** Bar diagram showing the relative abundance of the antimicrobial resistant drug classes across the 17 locations based on the read counts.

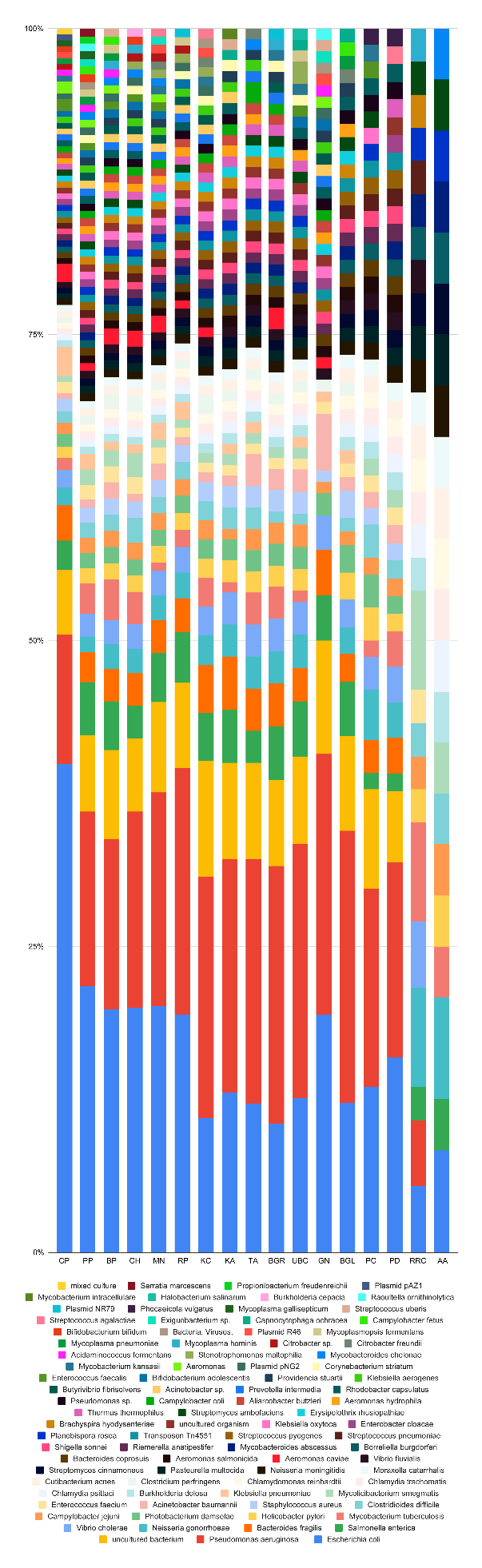

**Supplementary Figure 6:** Bar diagram showing the relative abundance of the hits of pathogens across the 17 locations.

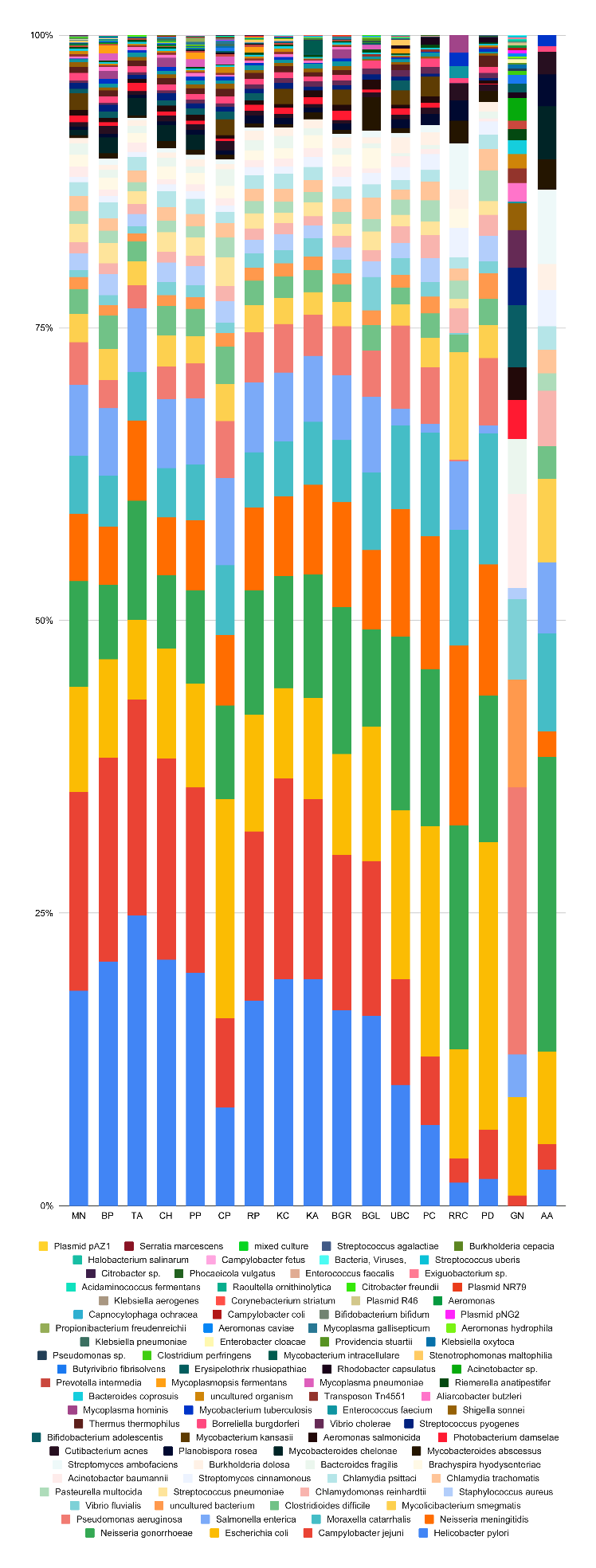

**Supplementary Figure 7:** Bar diagram showing the relative abundance of the pathogens across the 17 locations based on the read counts.

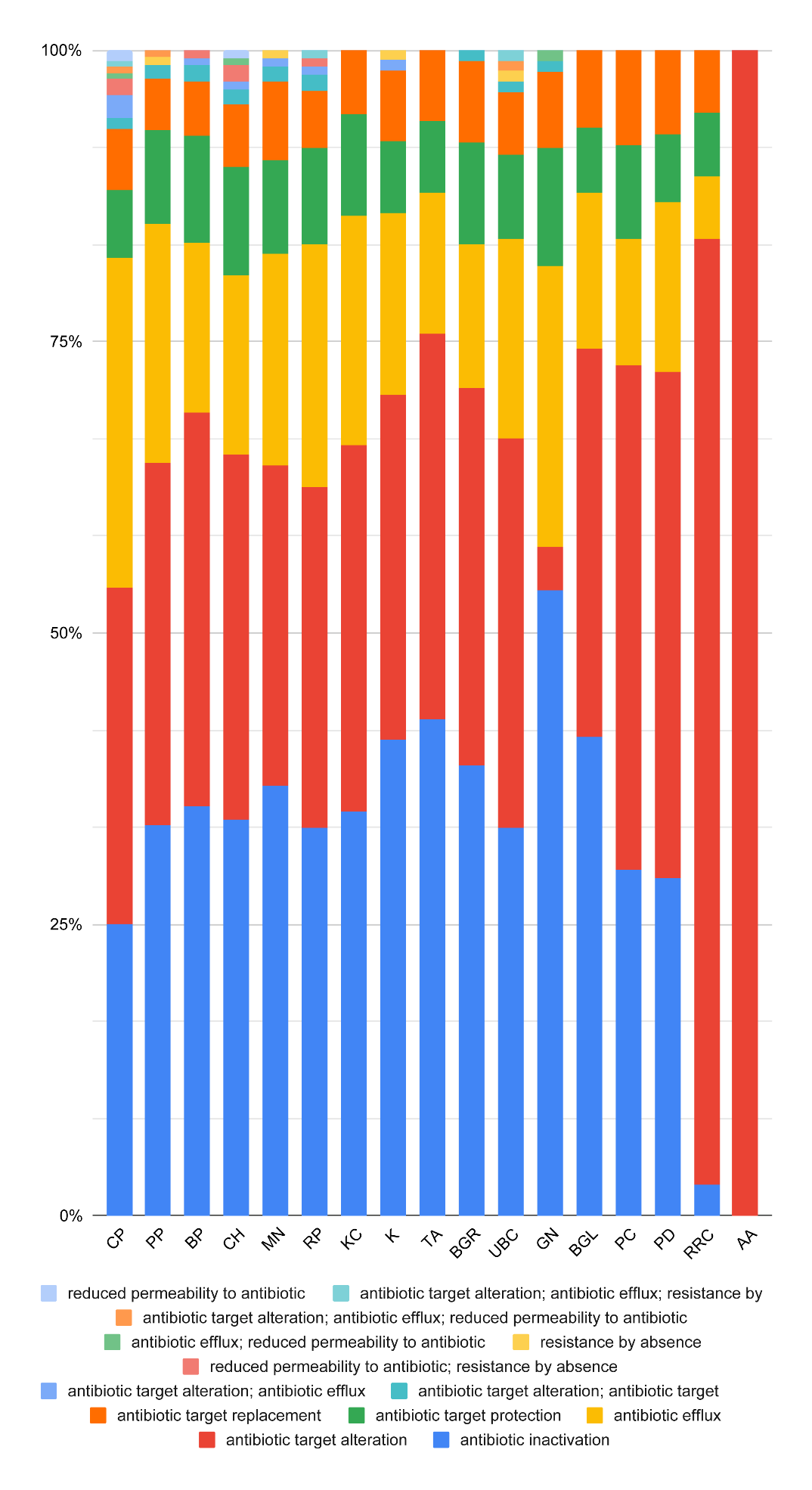

**Supplementary Figure 8:** Bar diagram showing the relative abundance of the hits of antimicrobial resistance mechanisms across the 17 locations.

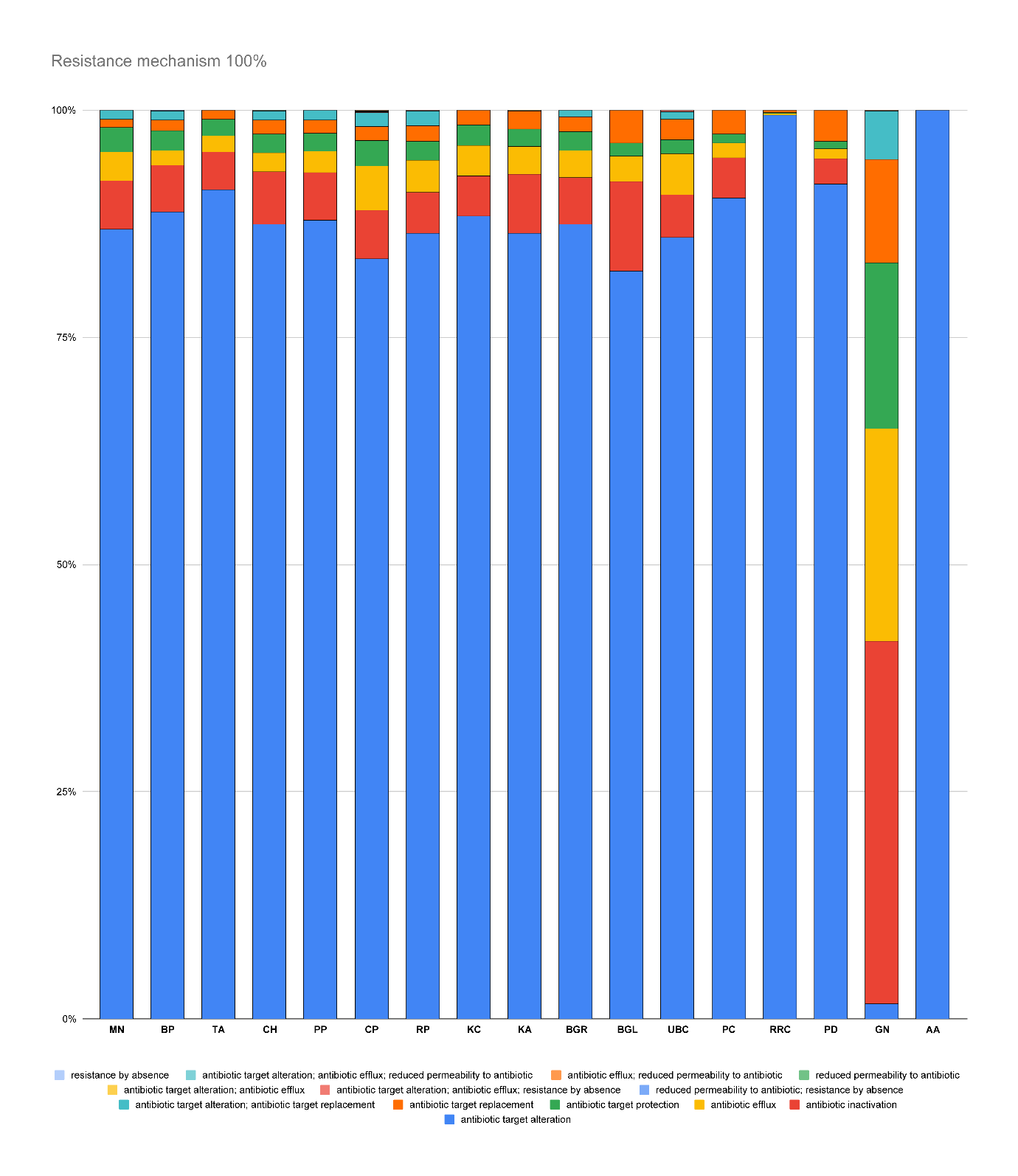

**Supplementary Figure 9:** Bar diagram showing the relative abundance of the antimicrobial resistance mechanisms across the 17 locations based on the read counts.

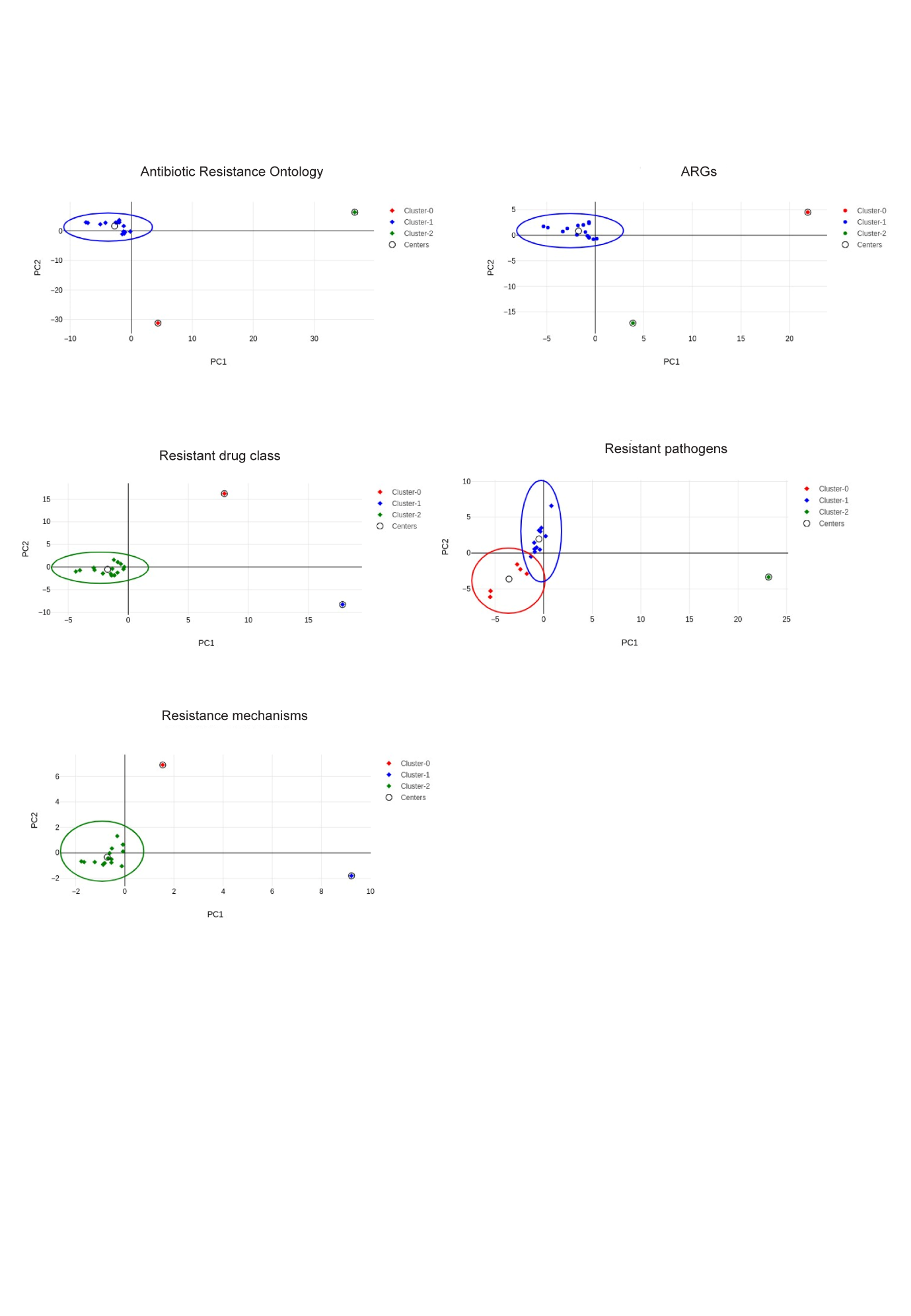

**Supplementary Figure 10: PCA plots showing the clustered locations**
